# The Invasive Respiratory Infection Surveillance (IRIS) Initiative reveals significant reductions in invasive bacterial infections during the COVID-19 pandemic

**DOI:** 10.1101/2020.11.18.20225029

**Authors:** Angela B Brueggemann, Melissa J Jansen van Rensburg, David Shaw, Noel McCarthy, Keith A Jolley, Martin CJ Maiden, Mark PG van der Linden, Zahin Amin-Chowdhury, Désirée E Bennett, Ray Borrow, Maria-Cristina C Brandileone, Karen Broughton, Ruth Campbell, Bin Cao, Carlo Casanova, Eun Hwa Choi, Yiu Wai Chu, Stephen A Clark, Heike Claus, Juliana Coelho, Mary Corcoran, Simon Cottrell, Robert J Cunney, Tine Dalby, Heather Davies, Linda de Gouveia, Ala-Eddine Deghmane, Walter Demczuk, Stefanie Desmet, Richard J Drew, Mignon du Plessis, Helga Erlendsdottir, Norman K Fry, Kurt Fuursted, Steve J Gray, Birgitta Henriques-Normark, Thomas Hale, Markus Hilty, Steen Hoffmann, Hilary Humphreys, Margaret Ip, Susanne Jacobsson, Jillian Johnston, Jana Kozakova, Karl G Kristinsson, Pavla Krizova, Alicja Kuch, Shamez N Ladhani, Thiên-Trí Lâm, Vera Lebedova, Laura Lindholm, David Litt, Irene Martin, Delphine Martiny, Wesley Mattheus, Martha McElligott, Mary Meehan, Susan Meiring, Paula Mölling, Eva Morfeldt, Julie Morgan, Robert M Mulhall, Carmen Muñoz-Almagro, David R Murdoch, Joy Murphy, Martin Musilek, Alexandre Mzabi, Amaresh Perez-Argüello, Monique Perrin, Malorie Perry, Alba Redin, Richard Roberts, Maria Roberts, Assaf Rokney, Merav Ron, Kevin Scott, Carmen L. Sheppard, Lotta Siira, Anna Skoczyńska, Monica Sloan, Hans-Christian Slotved, Andrew J Smith, Joon Young Song, Muhamed-Kheir Taha, Maija Toropainen, Dominic Tsang, Anni Vainio, Nina M van Sorge, Emmanuelle Varon, Jiri Vlach, Ulrich Vogel, Sandra Vohrnova, Anne von Gottberg, Rosemeire C Zanella, Fei Zhou

**Affiliations:** National Reference Centre for Streptococcus pneumoniae, University Hospitals Leuven, Leuven, Belgium; Department of Microbiology, Immunology and Transplantation, KU Leuven, Leuven, Belgium; National Reference Centre for Haemophilus influenzae, Laboratoires des Hôpitaux Universitaires de Bruxelles - Universitaire Laboratorium Brussel, Brussels, Belgium; Faculté de Médecine et Pharmacie, Université de Mons, Mons, Belgium; National Reference Centre for Neisseria meningitidis, Sciensano, Brussels, Belgium; National Laboratory for Meningitis and Pneumococcal Infections, Center of Bacteriology, Institute Adolfo Lutz, São Paulo, Brazil; National Microbiology Laboratory, Public Health Agency of Canada, Winnipeg, Manitoba, Canada; Department of Pulmonary and Critical Care Medicine, Center of Respiratory Medicine, National Clinical Research Center for Respiratory Diseases, Institute of Respiratory Medicine, Chinese Academy of Medical Sciences, Peking Union Medical College, Beijing, China; National Reference Laboratory for Haemophilus Infections, Centre for Epidemiology and Microbiology, National Institute of Public Health, Prague, Czech Republic; National Reference Laboratory for Meningococcal Infections, Centre for Epidemiology and Microbiology, National Institute of Public Health, Prague, Czech Republic; National Reference Laboratory for Streptococcal Infections, Centre for Epidemiology and Microbiology, National Institute of Public Health, Prague, Czech Republic; Department of Bacteria, Parasites and Fungi, Statens Serum Institut, Copenhagen, Denmark; Nuffield Department of Population Health, Big Data Institute, University of Oxford, Oxford, England, United Kingdom; Department of Zoology, University of Oxford, Oxford, England, United Kingdom; Blavatnik School of Government, University of Oxford, Oxford, England, United Kingdom; Immunisation and Countermeasures Division, National Infection Service, Public Health England, London, England, United Kingdom; Meningococcal Reference Unit, National Infection Service, Public Health England, Manchester Royal Infirmary, Manchester, United Kingdom; Public Health England, Immunisation and Countermeasures Division, London, United Kingdom; Respiratory and Vaccine Preventable Bacteria Reference Unit, National Infection Service, Public Health England, London, United Kingdom; Vaccine Preventable Bacteria Section, National Infection Service, Public Health England, London, England, United Kingdom; Warwick Medical School, University of Warwick, Warwick, England, United Kingdom; Finnish Institute for Health and Welfare (THL), Helsinki, Finland; Institut Pasteur, Invasive Bacterial Infections Unit and National Reference Centre for Meningococci and Haemophilus influenzae, Paris, France; Laboratory of Medical Biology and National Reference Centre for Pneumococci, Intercommunal Hospital of Créteil, Créteil, France; German National Reference Center for Streptococci, Department of Medical Microbiology, University Hospital RWTH Aachen, Aachen, Germany; German National Reference Center for Meningococci and Haemophilus influenzae, Institute for Hygiene and Microbiology, University of Würzburg, Würzburg, Germany; Microbiology Division, Public Health Laboratory Services Branch, Centre for Health Protection, Department of Health, HKSAR, PR China; Department of Microbiology, The Chinese University of Hong Kong, Hong Kong; Department of Clinical Microbiology, Landspitali, The National University Hospital of Iceland, Reykjavik, Iceland; Irish Meningitis and Sepsis Reference Laboratory, Children’s Health Ireland at Temple Street, Dublin, Ireland; Department of Clinical Microbiology, Royal College of Surgeons in Ireland, Beaumont Hospital, Dublin, Ireland; Department of Clinical Microbiology, Royal College of Surgeons in Ireland, Dublin, Ireland; Department of Clinical Microbiology, Beaumont Hospital, Dublin, Ireland; Government Central Laboratories, Ministry of Health, Jerusalem, Israel; Laboratoire National de Sante, Dudelange, Luxembourg; Department of Medical Microbiology and Infection Prevention and Netherlands Reference Laboratory for Bacterial Meningitis, Amsterdam University Medical Center, University of Amsterdam, Amsterdam, Netherlands; Meningococcal Reference Laboratory, Institute of Environmental Science and Research Limited, Porirua, New Zealand; Streptococcal Reference Laboratory, Institute of Environmental Science and Research Limited, Porirua, New Zealand; University of Otago, Christchurch, New Zealand; Public Health Agency, Belfast, Northern Ireland; National Reference Centre for Bacterial Meningitis, National Medicines Institute, Warsaw, Poland; Bacterial Respiratory Infection Service, Scottish Microbiology Reference Laboratories, Glasgow, Scotland, United Kingdom; Centre for Respiratory Diseases and Meningitis, National Institute for Communicable Diseases, Division of the National Health Laboratory Service, Johannesburg, South Africa; Division of Public Health Surveillance and Response, National Institute for Communicable Diseases, Division of the National Health Laboratory Service, Johannesburg, South Africa; Department of Pediatrics, Seoul National University College of Medicine, Seoul, South Korea; Division of Infectious Diseases, Department of Internal Medicine, Korea University Guro Hospital, Korea University College of Medicine, Seoul, South Korea; Instituto de Recerca Pediatrica, Hospital Sant Joan de Deu, Barcelona, Spain; Department of Microbiology, Tumor and Cell Biology, Karolinska Institutet, Stockholm, Sweden; Department of Clinical Microbiology, Karolinska University Hospital, Stockholm, Sweden; National Reference Laboratory for Neisseria meningitidis, Department of Laboratory Medicine, Clinical Microbiology, Faculty of Medicine and Health, Örebro University, Örebro, Sweden; Public Health Agency of Sweden, Solna, Sweden; Swiss National Reference Centre for invasive Pneumococci, Institute for Infectious Diseases, University of Bern, Bern, Switzerland; Public Health Wales, Cardiff, Wales, United Kingdom

## Abstract

**Background:** *Streptococcus pneumoniae, Haemophilus influenzae* and *Neisseria meningitidis* are leading causes of invasive diseases including bacteraemic pneumonia and meningitis, and of secondary infections post-viral respiratory disease. They are typically transmitted via respiratory droplets. We investigated rates of invasive disease due to these pathogens during the early phase of the COVID-19 pandemic.

**Methods:** Laboratories in 26 countries across six continents submitted data on cases of invasive disease due to *S pneumoniae, H influenzae* and *N meningitidis* from 1 January 2018 to 31 May 2020. Weekly cases in 2020 vs 2018-2019 were compared. *Streptococcus agalactiae* data were collected from nine laboratories for comparison to a non-respiratory pathogen. The stringency of COVID-19 containment measures was quantified by the Oxford COVID-19 Government Response Tracker. Changes in population movements were assessed by Google COVID-19 Community Mobility Reports. Interrupted time series modelling quantified changes in rates of invasive disease in 2020 relative to when containment measures were imposed.

**Findings:** All countries experienced a significant, sustained reduction in invasive diseases due to *S pneumoniae, H influenzae* and *N meningitidis*, but not *S agalactiae*, in early 2020, which coincided with the introduction of COVID-19 containment measures in each country. Similar impacts were observed across most countries despite differing stringency in COVID-19 control policies. There was no evidence of a specific effect due to enforced school closures.

**Interpretation:** The introduction of COVID-19 containment policies and public information campaigns likely reduced transmission of these bacterial respiratory pathogens, leading to a significant reduction in life-threatening invasive diseases in many countries worldwide.

## Research in context

### Evidence before this study

Invasive diseases caused by *S pneumoniae, H influenzae* and *N meningitidis*, and the risk of invasive bacterial disease subsequent to viral pneumonia, are well described in the scientific and medical literature. SARS-CoV-2 is a novel coronavirus that has led to a pandemic of disease called COVID-19 in 2020. Large-scale containment measures including school and workplace closures and complete national lockdowns have been implemented in most countries to prevent the transmission of SARS-CoV-2 and reduce the incidence of COVID-19. This widespread, global response to COVID-19 is unprecedented and may have led to a reduction in transmission of other respiratory pathogens. We searched PubMed, bioRxiv and medRxiv for articles published in English up to 31 December 2019, prior to the COVID-19 pandemic, using search terms ‘pandemic’ AND ‘microbial transmission’ (or ‘transmission’) AND ‘containment’. 262 papers were identified. Although strategies for containment and reducing transmission of an epidemic or pandemic pathogen have been well described in the literature, there is currently an evidence gap on the extent to which large-scale containment measures implemented during a pandemic reduce the burden of infectious diseases due to pathogens other than the one causing the pandemic.

### Added value of this study

Since invasive diseases result in high morbidity and mortality, many countries have a statutory duty to notify health authorities on suspicion of infection and request referral of any isolates to microbiology reference laboratories for surveillance purposes. We used existing laboratory data to address the impact of COVID-19 and associated containment measures on invasive diseases caused by *S pneumoniae, H influenzae, N meningitidis* and *Streptococcus agalactiae* (the latter as a comparator organism). We rapidly established an international network of laboratories in 26 countries, compiled a large invasive disease dataset of >80,500 case records, analysed data on national policy decisions and containment measures in each country, and examined the movements of people during the early months of the COVID-19 pandemic. This demonstrated that invasive disease due to *S pneumoniae, H influenzae* and *N meningitidis* plummeted in all participating countries with the introduction of COVID-19 containment measures in early 2020. The decreases in invasive disease were largely consistent across countries, despite variation in the stringency of containment measures adopted in the early stages of the COVID-19 pandemic.

### Implications of all the available evidence

High-quality, prospective microbiological disease surveillance is crucial to global health. COVID-19 containment measures beneficially reduce transmission of respiratory pathogens and associated diseases, but they also impose a heavy burden on society that must be carefully considered before going forward. Invasive diseases due to *S pneumoniae, H influenzae* and *N meningitidis* are among the leading causes of death and disability worldwide. Safe and effective vaccines for all three pathogens are used in many, but not all, countries and should be implemented more widely. Finally, the invasive disease burden is likely to increase as COVID-19 containment measures are decreased and therefore ongoing microbiological surveillance such as that demonstrated in this study is essential.

## Introduction

Invasive bacterial diseases, in particular bacteraemic pneumonia, meningitis and septicaemia, are leading causes of global morbidity and mortality among all age groups but especially among young children, adolescents and older adults. The most common causes of these life-threatening diseases are *Streptococcus pneumoniae* (pneumococcus), *Haemophilus influenzae*, and *Neisseria meningitidis* (meningococcus), which normally reside in the nasopharynx or throat of healthy individuals and are transmitted person-to-person via the respiratory route (1-4).

Worldwide in 2016, there were 336 million episodes of lower respiratory infections that led to 2.4 million deaths. Respiratory infections were the sixth leading cause of death among all ages and the most common cause of death in children less than five years old. *S pneumoniae* was estimated to have caused 197 million episodes of pneumonia that led to over 1.1 million deaths, more deaths than all other aetiologies combined (5). Globally, the number of deaths due to meningitis among all ages was around 300,000, from an estimated 2.8 million meningitis episodes. Meningitis outbreaks due to these three bacteria, and *N meningitidis* in particular, have occurred all over the world (5, 6).

SARS-CoV-2 is a novel coronavirus first recognised as a cause of respiratory infection in early 2020 and it causes COVID-19 disease in humans. 34 million COVID-19 cases and 1 million COVID-19 related deaths were reported worldwide as of 30 September 2020 (7). Viral respiratory infections are associated with an increased risk of subsequent bacterial infections, especially invasive diseases and pneumonia, eg, the high mortality of the 1918 influenza pandemic was strongly associated with post-viral pneumonia caused by *S pneumoniae* and no antimicrobials to treat bacterial pneumonia (8-12). Therefore, there is the potential for increased rates of invasive bacterial diseases subsequent to SARS-CoV-2 infection. Alternatively, containment measures initiated in many countries to reduce viral transmission may result in decreased invasive disease due to a concomitant reduction in transmission of respiratory-associated bacteria.

Given the severity of the diseases they cause, clinical microbiology laboratories in many countries are required to report invasive infections due to *S pneumoniae, H influenzae* and *N meningitidis* to national health authorities and many also request referral of any isolates to reference laboratories for surveillance purposes. We established the Invasive Respiratory Infection Surveillance (IRIS) Initiative with a global network of reference laboratories in 26 countries, to rapidly analyse laboratory-confirmed invasive bacterial infection data during the COVID-19 pandemic, as compared to rates of invasive disease in previous years.

We show that in early 2020 the incidence rates of invasive disease due to *S pneumoniae, H influenzae* and *N meningitidis* plummeted in every country in the IRIS network, relative to rates of disease in 2018 and 2019. These decreases were strongly correlated with the timing of government responses to COVID-19, as measured by the Oxford COVID-19 Government Response Tracker (OxCGRT), and changes in the movement of people as measured by Google COVID-19 Community Mobility Reports (Google CCMR) (13, 14). In contrast, *S agalactiae* is not transmitted via the respiratory route, and the incidence rates of invasive disease due to *S agalactiae* in nine IRIS laboratories in 2020 were no different to the two previous years. The IRIS Initiative rapidly established a peer-to-peer international network of laboratories to monitor changes in invasive bacterial diseases. Our findings are consistent with a dramatic reduction in life-threatening invasive diseases resulting from reduced person-to-person transmission of three major bacterial respiratory pathogens.

## Methods

Microbiology laboratories with established invasive disease surveillance systems were approached to join the IRIS network. All IRIS laboratories were national reference laboratories apart from the laboratory in Spain (which represented Catalonia) and China (which represented one Institute in Beijing). Confirmed cases of invasive disease due to *S pneumoniae, H influenzae* or *N meningitidis* (ie, detected from a normally sterile site and/or from a patient with invasive disease) plus the sampling date were collected. All laboratories submitted data for *S pneumoniae* and the majority submitted data for *H influenzae* and *N meningitidis*. Nine laboratories submitted *S agalactiae* (Lancefield group B streptococcus) data as a control pathogen. *S agalactiae* is not a respiratory pathogen but is often found in the healthy gastrointestinal and lower genital tract and is a risk factor for invasive disease in pregnant women and neonates (15). Neonatal disease is attributable to vertical transmission from mother to baby during childbirth; however, invasive disease due to *S agalactiae* is increasing among non-pregnant adults. The mode of transmission among adults is less clear (16).

No patient-identifiable data were collected. A private IRIS dataview was set up as part of the PubMLST suite of databases and IRIS participants were able to upload and view study data (17). A subset of the French *N meningitidis* dataset was published previously (18). Cases diagnosed between 1 January 2018 and 31 May 2020 were analysed. Case counts were summed by the International Organization for Standardization (ISO) week.

The OxCGRT collects information on policies and interventions that governments have taken during the COVID-19 pandemic (13). Data are collected from >180 countries for 18 different indicators, which are combined into composite indices that measure the magnitude of government responses. The Stringency Index, which combines data on public information campaigns and eight physical distancing measures, was used in these analyses. The OxCGRT dataset was downloaded on 5 October 2020, the daily Stringency Index data were converted to a mean ISO week value and merged with the IRIS case data.

The Google CCMR provide anonymised, aggregated within-country data on the movement of people by capturing mobile device location history data from Google users in six categories (grocery/pharmacy, parks, transit stations, retail/recreation, residential, and workplaces) (14). Data that could identify individuals are not available. Daily mobility data were calculated as a percentage change from the baseline day, the median value from the 5-week period from 3 January to 6 February 2020. Google CCMR data were downloaded on 5 October 2020.

Statistical evidence for the impact of an interruption due to the COVID-19 response was assessed using generalised linear models, and both the step and linear slope changes following this interruption were estimated. For *S pneumoniae*, models were: (i) individually fitted to the dataset of each country, utilising a Poisson distribution and including a scaling factor to correct for overdispersion, and results were summarised by meta-analysis; and (ii) fitted as a single mixed effects negative binomial model. Country-specific interruption time points were based on Google CCMR data for workplace mobility, selecting the week containing the midpoint of the decline in work-associated mobility. No Google CCMR data were available for Iceland and China, therefore, Iceland was assigned the modal week of other European countries, while China’s interruption point was based on policy/news releases and set at week five (Supplementary Table 1).

In the individual country models, Fourier sequences with two terms were fitted to the model for each country to account for seasonal variation in case numbers, and a linear term for long term trend. This produced incident rate ratios, standard errors and confidence intervals for each country for the step change: the ratio of cases per week after the interruption point compared to the number of cases that occurred before interruption, and slope change for further ongoing decline from this time point. A meta-analysis of the estimates for each country was performed to generate pooled effect sizes, using the inverse-variant fixed effects and restricted maximum likelihood random effects method. This assessed the effect of the step, the slope, and linear combinations of these, to estimate the change from expected numbers of *S pneumoniae* cases over 4- and 8-week periods following the interruption time point. The mixed effects model included one sine and cosine term for seasonality (a second term was unsupported), a linear effect in time, and a step and slope for country-specific interruption time in the fixed effects part of the model. Week numbers were edited for Southern Hemisphere countries (ie, running from week 27 to 26) to allow joint modelling of seasonality. Random effects were included for seasonality, step and slope variables. Widespread school closures (OxCGRT level 2 or 3) were used to model the impact of specific policy changes alongside general changes in behaviours indexed by the Google CCMR workplace mobility data. A further country-specific interruption was modelled using the date of school closures and allowing both a step and slope change from that date. Models were individually checked for assumption violation. Sensitivity testing was performed to assess whether a lag produced better model fit.

Models were also run on the combined weekly count data for each of the four pathogens. These analyses included those countries that had submitted data for all of *S pneumoniae, H influenzae* and *N meningitidis* to allow comparability. A common interruption week (week 11, when the COVID-19 pandemic was declared) was applied and step and slope parameters were estimated. A counterfactual estimate was calculated for the expectation of cases in the absence of an interruption to the time series. Statistical analyses were performed in Stata 16.1 and R version 3.6.1.

## Results

### Overall reduction in invasive diseases due to *S pneumoniae, H influenzae* and *N meningitidis*

Twenty-six laboratories submitted invasive disease data to IRIS for one or more pathogens: *S pneumoniae* (26 countries; 62,837 cases); *H influenzae* (24 countries; 7,796 cases); *N meningitidis* (21 countries; 5,877 cases). Weekly data were analysed to assess changes in the numbers of cases in 2020 versus the previous two years. There was a substantial and sustained reduction in the number of invasive cases of *S pneumoniae, H influenzae* and *N meningitidis* diagnosed between March and May 2020 (Figure 1). This was in clear contrast to 2018-2019, when the overall numbers of *S pneumoniae* and *H influenzae* cases were very similar each year. A comparable reduction was observed for *N meningitidis* in 2020, although the number of cases in 2018 vs 2019 varied (2,700 vs 2,457, respectively). The reductions in the weeks following the interruption to the expected time series were strongly supported statistically (P <0.0001) from likelihood ratio tests of models with and without parameters for the reduction (Supplementary Figure 1).

**Figure 1.**
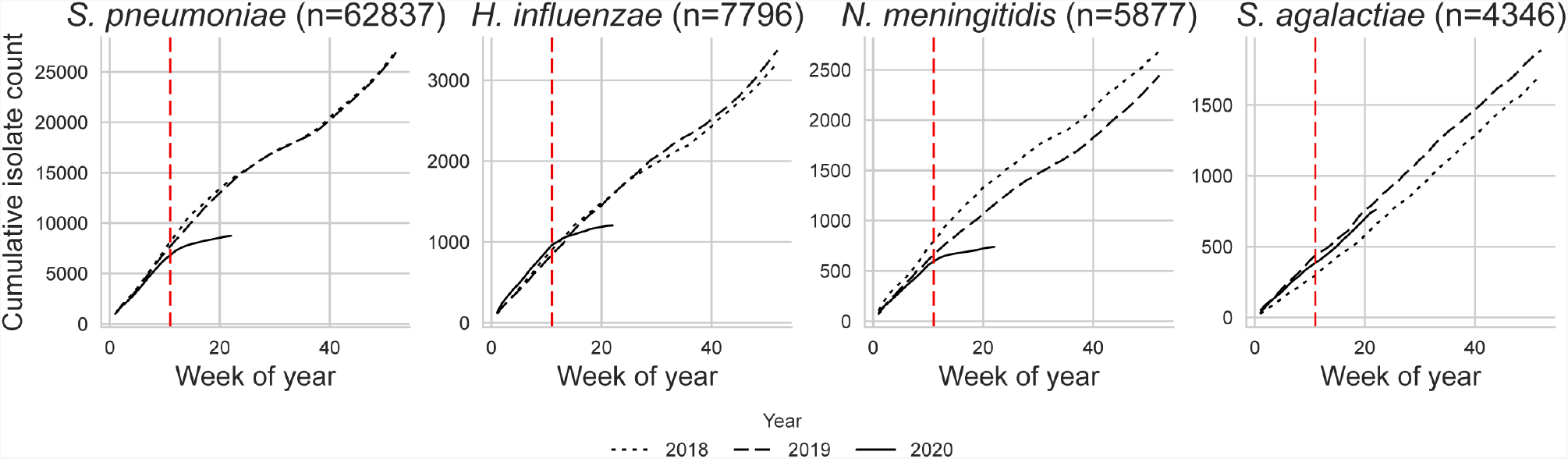
Cumulative curves depicting the number of cases collected by IRIS laboratories each week from 1 January 2018 through 31 May 2020. The World Health Organization (WHO) officially declared the COVID-19 pandemic in week 11 of 2020 (red hashed line).

### No change in *S agalactiae* submissions during the same period

One explanation for the reduction in numbers of invasive cases was that routine invasive disease surveillance was disrupted whilst countries were responding to COVID-19; however, IRIS laboratories did not observe significant disruptions in routine submissions. To test this further, we analysed 4,346 cases of *S agalactiae* submitted over the same time period from nine IRIS laboratories. There was no evidence of any change in *S agalactiae* submissions in 2020 versus 2018-2019, supporting the view that the reductions in *S pneumoniae, H influenzae* and *N meningitidis* cases in 2020 were reflective of actual decreases in disease incidence and not a consequence of disruptions in routine case reporting or isolate referral (Figure 1).

### Reductions in invasive disease relative to the stringency of COVID-19 containment measures

To assess the effect of COVID-19 containment measures on the reduction in invasive infections, the weekly bacterial case submission data were compared to the OxCGRT Stringency Index calculated for each country. The Stringency Index provided a combined estimate of the stringency of public information campaigns plus containment measures including school closures, workplace closures, cancellation of public events, restrictions on gatherings, closures of public transport, stay at home requirements, restrictions on internal movement and international travel controls. The results of these analyses for *S pneumoniae* are depicted in Figure 2 and the data for *H influenzae, N meningitidis* and *S agalactiae* are provided in Supplementary Figures 2-4.

**Figure 2.**
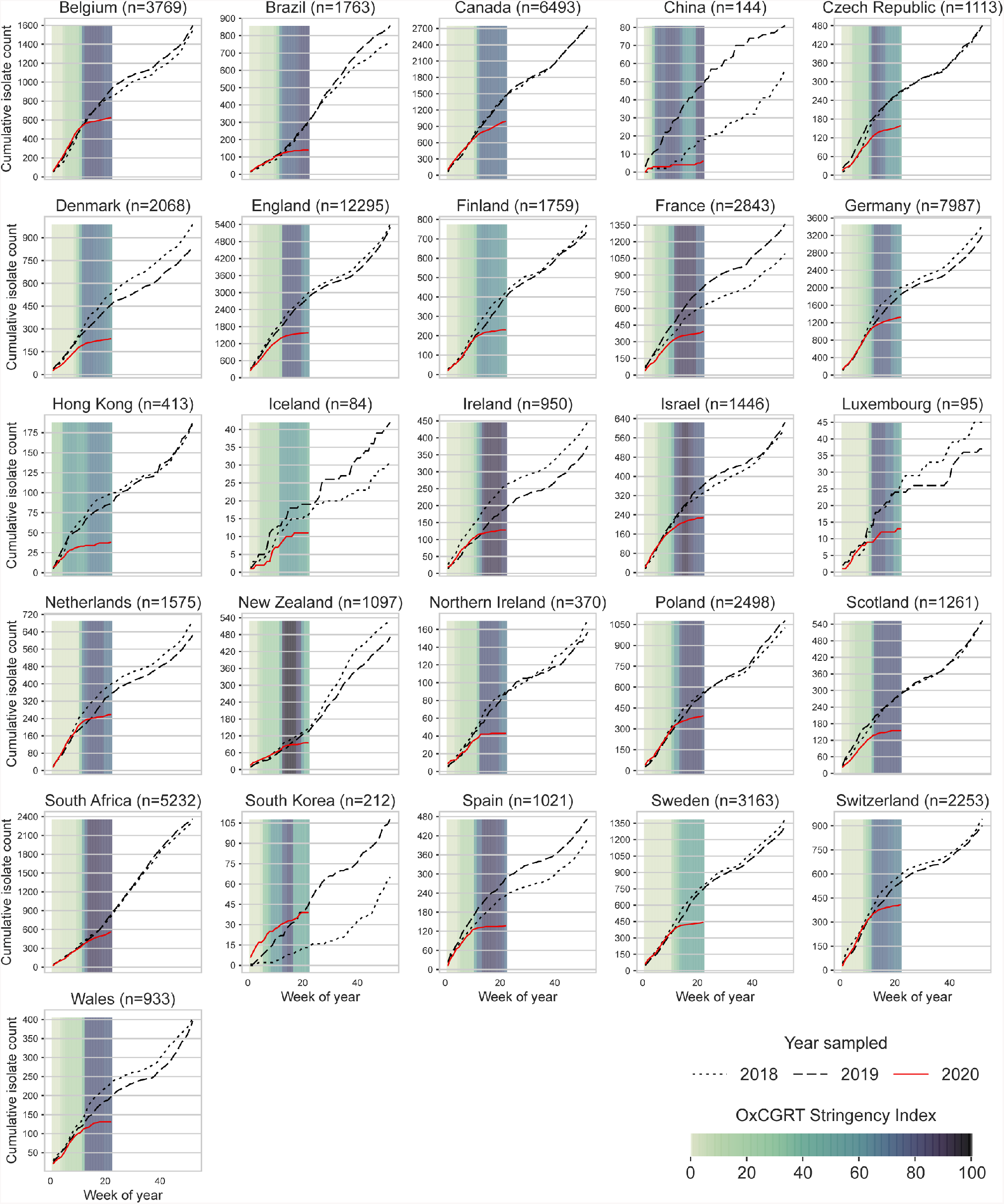
Annual cumulative curves of invasive S pneumoniae cases submitted to IRIS laboratories in 26 countries from 1 January 2018 through 31 May 2020. Coloured bars represent the mean weekly OxCGRT Stringency Index values on a scale from 0-100. Larger (darker) values indicate that higher stringency measures were enacted within a country. Note that data from South Korea were submitted from two surveillance networks, one of which started invasive disease surveillance in September 2018, so data presented here for that hospital are only from September 2018 onwards, whereas the data from the other hospital are from January 2018 onwards.

The WHO officially declared the COVID-19 pandemic in week 11 of 2020 and by that point all countries represented in IRIS had initiated some COVID-19 responses, ranging from 11-81 on the Stringency Index scale (Figure 2). By week 15, countries had rapidly increased containment measures and public information campaigns: fifteen countries had a Stringency Index score >80, eight countries (Brazil, Canada, China, Czech Republic, Denmark, Germany, Hong Kong and Switzerland) had scores of 60-80, and three countries (Finland, Iceland and Sweden) had scores of 45-60. China initiated stringent containment measures and public information campaigns in week 4, then reduced the measures slightly and by week 20 increased to high stringency measures. Whilst the stringency of the imposed containment measures varied by country, there was a dramatic reduction in *S pneumoniae* invasive disease notifications among all IRIS participants and this reduction was sustained through the end of May. Similar trends were observed for *H influenzae* and *N meningitidis* but not *S agalactiae* (Supplementary Figures 2-4).

### Changes in human behaviour also lead to a reduction in invasive disease

To assess the extent to which societal choices also influenced the reductions in invasive disease, we compared the movement of people within each country using the Google CCMR data (Figure 3). All 26 countries, regardless of the stringency of the containment measures imposed by their governments, experienced a decrease in workplace visits and an increase in time spent in residential areas. The largest changes in movement (as compared to baseline) occurred at the time the WHO declared the COVID-19 pandemic and shortly thereafter.

**Figure 3.**
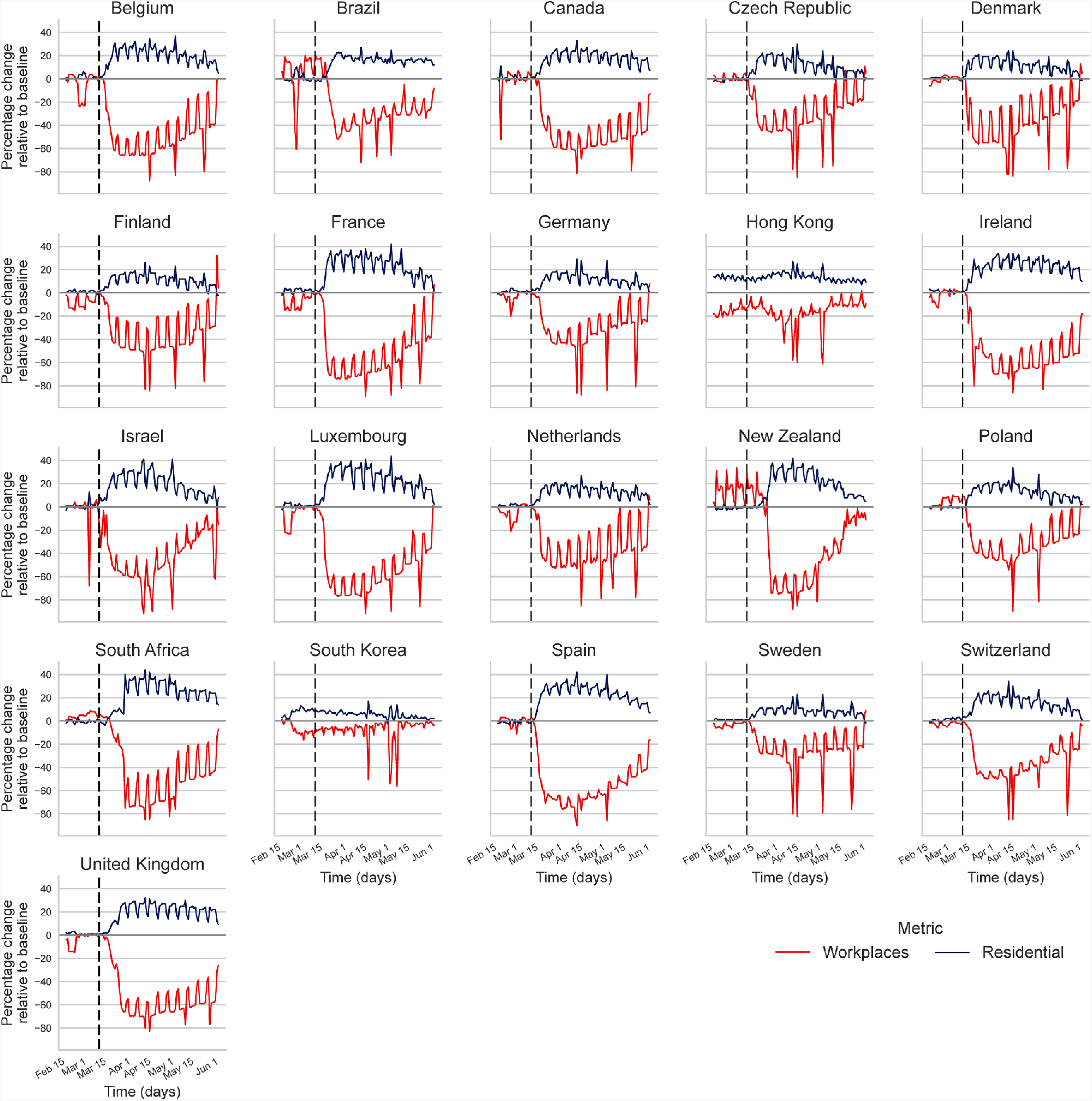
Assessment of the movement of people in IRIS countries using Google CCMR data. Workplaces and Residential data are plotted for each country participating in IRIS except for Iceland, due to the potential for privacy breaches related to the small population size. Data from China were not collected. Google CCMR data for the United Kingdom aggregated England, Scotland, Wales, and Northern Ireland as one dataset. The vertical line marks week 11, when the WHO officially declared the COVID-19 pandemic. The periodic nature of the graphs is because these are daily data and the movement of people changes at the weekends; the sharp data spikes typically represent national holidays such as the Easter weekend in mid-April.

Countries that experienced the most stringent COVID-19 containment measures also experienced the largest changes in movements around workplaces and residences, but even those countries with moderate containment measures (ie, Finland and Sweden) experienced large changes in population movements in the same positive or negative direction as all other countries. Changes in the movement of people within Hong Kong and South Korea were less variable and occurred earlier. Countries in the southern hemisphere (Brazil, New Zealand, and South Africa) had different patterns of movement in the early weeks of 2020, in part because of summer holidays. By the end of May 2020, the mobility data in many countries were shifting back to baseline levels and movements around workplaces were beginning to increase above baseline in several countries; however, Brazil, Canada, Ireland, South Africa, Spain, and the United Kingdom had not yet reverted back to baseline levels of movement for either workplaces or residences.

### The timing of containment measures and changes in the movements of people are significantly associated with reduced invasive pneumococcal disease

A total of 62,837 cases of *S pneumoniae* invasive disease were reported between 1 January 2018 and 31 May (week 22) of 2020. The models estimated that social changes caused by the COVID-19 pandemic led to a 38% decrease in the incidence rate of reported *S pneumoniae* invasive infections (IRR 0.62, 95% CI 0.55-0.70) immediately following the interruption (step change parameter) followed by an additional 13% average weekly reduction during the following weeks (IRR 0.87, 95% CI 0.84-0.89). Similar results were obtained from the combined model analysis for both step change (IRR 0.56, 95% CI 0.49-0.65) and slope (IRR 0.87, 95% CI 0.84-0.90) parameters and there was no strong evidence to favour a lag between movement changes and effects on infection rates. Although there was some variation among countries on the individual country analyses, the effect sizes were largely similar (Figure 4).

**Figure 4.**
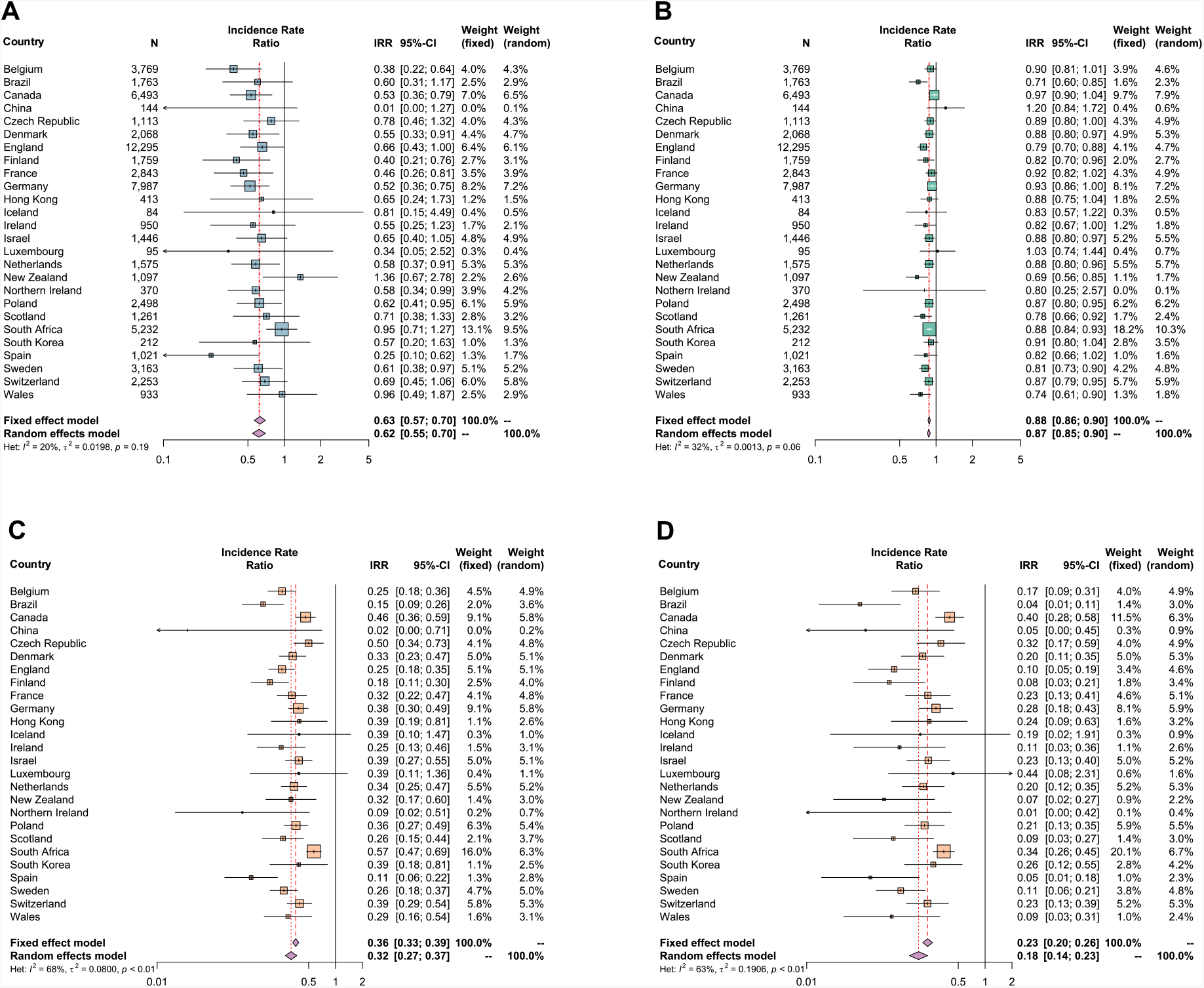
Forest plots depicting the results of interrupted time series models applied to the S pneumoniae data. The first two panels depict the results of a model that allows for a step (panel A) or slope (panel B) reduction in invasive disease. Panels C and D depict results of a model that combines step and slope, and plots the country-specific incidence rate ratios after four weeks (panel C) or eight weeks (panel D) from the point at which national mobility was significantly interrupted (see Supplementary Table 1 for weeks of interruption).

Overall, as compared to expectations based on the 2018 and 2019 time series, this translated into a decrease in the incidence rate of reported *S pneumoniae* infections of 68% at 4 weeks (IRR 0.32, 95% CI 0.27-0.37) and 82% at 8 weeks (IRR 0.18, 95% CI 0.14-0.23) following the week when movement changes were noted. Adding country-specific terms for school closures into the combined model analysis did not substantially improve fit (p=0.092) with strong support remaining for a decrease following reduced mobility (as indexed by Google CCMR data) and little evidence for additional effects linked specifically to school closures.

## Conclusions

The COVID-19 pandemic has been an unprecedented global situation. Governments have imposed containment measures in an attempt to protect their communities, slow the transmission of SARS-CoV-2, and reduce morbidity and mortality due to COVID-19. Healthcare systems in many high-income countries have been stretched beyond what had previously thought possible.

These IRIS data demonstrated a significant reduction in invasive diseases caused by three bacterial pathogens that are, like SARS-CoV-2, transmitted via the respiratory route. The most plausible and parsimonious explanation for this was the interruption of person-to-person bacterial respiratory transmission. Despite wide variations in stringency, the timing of containment measures imposed in all countries represented here coincided with this rapid reduction in invasive diseases. The mobility data suggested that people also voluntarily reduced their personal risks during the early stages of the pandemic. We therefore contend that the IRIS data were a proxy for the effectiveness of public health measures undertaken to reduce respiratory pathogen transmissions. It should not be assumed that the effect on bacterial transmission was identical to that of SARS-CoV-2, but it is likely to follow a similar trajectory.

Since the ecological niche of *S pneumoniae* is typically the paediatric nasopharynx, the extent to which school closures explained the significant reduction in invasive diseases caused by *S pneumoniae* was assessed. Adding parameters based on the week of enforced school closure did not offer a significantly improved fit over the model with parameters for workplace mobility data as a general measure of changed behaviours. Therefore, whilst school closures would have contributed to the observed reductions in movement of people and reduced transmission of *S pneumoniae*, in addition to physical distancing and other measures, closing schools did not produce a detectable additional reduction in invasive pneumococcal disease in these data. It is also possible that shielding older adults regardless of school closures may have limited bacterial transmission (19-21).

Globally, morbidity and mortality rates associated with *S pneumoniae, H influenzae* and *N meningitidis* are high. Safe and effective vaccines are available, and whilst these vaccines do not protect against all serotypes of each pathogen, they have been successfully implemented in childhood immunisation programmes of many countries. Nevertheless, vaccination is nowhere near comprehensive and public health efforts must remain focused on these three pathogens (22-25). Furthermore, in the context of preventing transmission, the current *S pneumoniae, H influenzae* serotype b and *N meningitidis* conjugate-polysaccharide vaccines are successful in large part because they can reduce bacterial colonisation and subsequent transmission, and these IRIS data underline the importance of reducing person-to-person transmission of respiratory pathogens (26).

The IRIS study highlights the crucial importance of active microbiological surveillance for public health and the fundamental role that reference laboratories play in this. Surveillance is most effective when it is performed consistently, provides high-quality data, and continues uninterrupted for many years so that emerging trends can be detected with confidence. Here, we have compared the previous two years to 2020 because the rates of disease were so obviously altered in 2020, but most IRIS laboratories have been undertaking high quality surveillance for many years.

The IRIS Initiative was quickly established through existing international public health/academic networks in response to COVID-19, and the network of collaborators and the data they produce will be an invaluable resource for observing and investigating future changes. Additional disease perturbations are to be expected as COVID-19 related containment measures are modified, which will allow greater insight into the impact of specific public health interventions. The IRIS laboratories will play a central role in rapidly detecting further invasive disease perturbations in each of their countries.

Potential limitations in these analyses include the completeness of the submissions to individual laboratories, which would mitigate the observed reduction in invasive diseases. However, in many countries these organisms are legally notifiable and these submissions are made routinely by well-established systems, often to guide diagnosis or public health action, all of which continued throughout this period. Furthermore, the *S agalactiae* submissions over the same period were as expected. It is also difficult to account for changes that could reduce microbial transmission but that were less easily measured, eg, parents keeping their children out of school from a fear of coronavirus infection but before schools were officially closed. This would have the effect of underestimating the impact of containment measures on reductions in disease transmission.

The strengths of the IRIS Initiative included the establishment of a network of national laboratories and surveillance programmes, which rapidly provided high quality invasive bacterial disease data from experienced laboratories. The IRIS network included 26 countries across six continents and a large overall dataset. The consistency in the trends observed across all individual datasets provided confidence in the data interpretations. Uniquely, we analysed invasive bacterial disease data, OxCGRT indices and Google CCMR data per country, which provided the means to assess the associations between country-specific containment measures, changes in the movements of human populations and the corresponding reductions in invasive diseases.

SARS-CoV-2 has been a stark reminder that infectious diseases remain a major threat to the lives and livelihoods of people across the globe. The COVID-19 pandemic has also revealed opportunities to accelerate advances in medicine and technology. Whilst COVID-19 is the current infectious disease challenge, others remain and there will be emerging pathogens in the future. This study provides an example of how the international public health and scientific community can collaborate to apply knowledge, experience and data analyses to improve global health.

## Data Availability

A list of anonymised cases with sampling dates is available upon request.
OxCGRT data are publicly available and found here: https://covidtracker.bsg.ox.ac.uk/
Google Mobility Reports data are publicly available and found here: https://www.google.com/covid19/mobility/

## Authors’ Contributions

ABB, MJvR, KAJ, MCJM and MvdL conceived of the IRIS Initiative, set up the database infrastructure and recruited laboratories to join IRIS. All authors were involved in the acquisition and/or processing of microbiological, OxCGRT and/or Google CCMR data. ABB, MJvR, DS and NM analysed and interpretated the overall data. ABB, MJvR, DS and NM wrote the first draft of the paper. All authors reviewed and revised the paper critically for important intellectual content and approved the final version to be published. All authors agreed to be accountable for all aspects of the work.

## Acknowledgments

The authors would like to acknowledge the assistance of laboratory personnel who made important contributions to the microbiological data in this study: Wendy Keijzers, Ilse Schuuman and Agaath Arends in the Netherlands Reference Laboratory for Bacterial Meningitis, Amsterdam, The Netherlands; Marie-Cécile Ploy, Carole Grelaud and all the microbiologists of the French Regional Observatory of Pneumococci Network; Alice Enefer, Anna Lewis, Karina Micah, Samuel Rose, Chenchal Dhami and Roger Daniel at the Public Health England Respiratory and Vaccine Preventable Bacteria Unit, London, England; Xilian Bai, Aiswarya Lekshmi, Jay Lucidarme, Andrew Walker, Lloyd Walsh and Laura Willerton at the Public Health England Meningococcal Reference Unit in Manchester, England; and Ana Paula Silva de Lemos e Samanta Cristine Grassi Almeida at the National Laboratory for Meningitis and Pneumococcal Infections, São Paulo, Brazil.

## Funding

The infrastructure for the IRIS initiative was funded by a Wellcome Trust Investigator Award to ABB (grant number 206394/Z/17/Z) and a Wellcome Trust Biomedical Resource Grant to MJCM, ABB and KAJ (grant number 218205/Z/19/Z). The German Reference Center for Streptococci receives financial support from Pfizer and Merck. The German National Reference Laboratory for Meningococci and *Haemophilus influenzae* is supported by the Robert Koch Institute with funds from the Federal Ministry of Health (funding code 1369-237). The Irish Meningitis and Sepsis Reference Laboratory has received support from Children’s Health Ireland at Temple Street, the Royal College of Surgeons in Ireland, Health Protection Surveillance Centre and the SpID-Net project, which has received funding from the European Centre for Disease Prevention and Control (ECDC) and Horizon 2020. MC has previously received a professional fee from Pfizer (Ireland), an unrestricted research grant from Pfizer Ireland (2007-2016) and an Investigator Initiated Reward from Pfizer Ireland in 2018 (W1243730). HH has received research funds from Astellas and Pfizer and a professional fee from Pfizer. The Polish data collection was partially supported by the Ministry of Health within the framework of the National Programme of Antibiotic Protection (NPOA) and by the Ministry of Science and Higher Education (Mikrobank 2 Programme). Funding for materials, equipment and human resources associated with the laboratory in Catalonia, Spain was provided by Agencia de Salut Pública de Catalunya and Sant Joan de Deu Foundation. BHN has received funding from the Knut and Alice Wallenberg Foundation, the Swedish Research Council and Region Stockholm. The funders had no role in the collection, analysis, interpretation of data or in the writing of and decision to submit the article for publication.

## Notes

### Competing Interest Statement

The German Reference Center for Streptococci receives financial support from Pfizer and Merck. MC has previously received a professional fee from Pfizer (Ireland), an unrestricted research grant from Pfizer Ireland (2007-2016) and an Investigator Initiated Reward from Pfizer Ireland in 2018 (W1243730). HH has received research funds from Astellas and Pfizer and a professional fee from Pfizer. The funders had no role in the collection, analysis, interpretation of data or in the writing of and decision to submit the article for publication.

### Author Declarations

Prof Kevin Marsh is the chair of OxTREC, the University of Oxford ethical committee that considers health related research conducted by Oxford-associated researchers where the research is primarily conducted outside of the UK. He confirmed that OxTREC would not require this study to go through ethical review, because it involves routine surveillance and no possibility of individual patients being identified. Prof Marsh also stated that this is the view that is taken by other ethical committees that he is familiar with. https://researchsupport.admin.ox.ac.uk/governance/ethics/committees/oxtrec

