## Supplementary table and figures for "The Invasive Respiratory Infection Surveillance (IRIS) Initiative reveals significant reductions in invasive bacterial infections during the COVID-19 pandemic"

**Brueggemann AB et al.**

**Supplementary Table 1. Country-specific weekly interruption time points used in the time series analyses.**

| Country | Week of the year 2020 |
|---|---|
| Belgium | 12 |
| Brazil | 12 |
| Canada | 12 |
| China | 5 |
| Czech Republic | 11 |
| Denmark | 11 |
| England | 12 |
| Finland | 12 |
| France | 11 |
| Germany | 12 |
| Hong Kong | 7 |
| Iceland | 11 |
| Ireland | 12 |
| Israel | 11 |
| Luxembourg | 11 |
| Netherlands | 11 |
| New Zealand | 13 |
| Northern Ireland | 12 |
| Poland | 11 |
| Scotland | 12 |
| South Africa | 12 |
| South Korea | 8 |
| Spain | 11 |
| Sweden | 11 |
| Switzerland | 11 |
| Wales | 12 |

*Note: The chosen week for each country was based on the Google COVID-19 Community Mobility Reports data (see Figure 3 in the main text), selecting the week containing the midpoint of the decline in work-associated mobility. No Google data were available for Iceland (the small national population presents a possible privacy breach) and China (due to censorship of Google data). Iceland was assigned the modal week of other European countries, while China's interruption point was based on policy and news releases and set at week 5 of 2020.*

**Supplementary Figures 1-4**

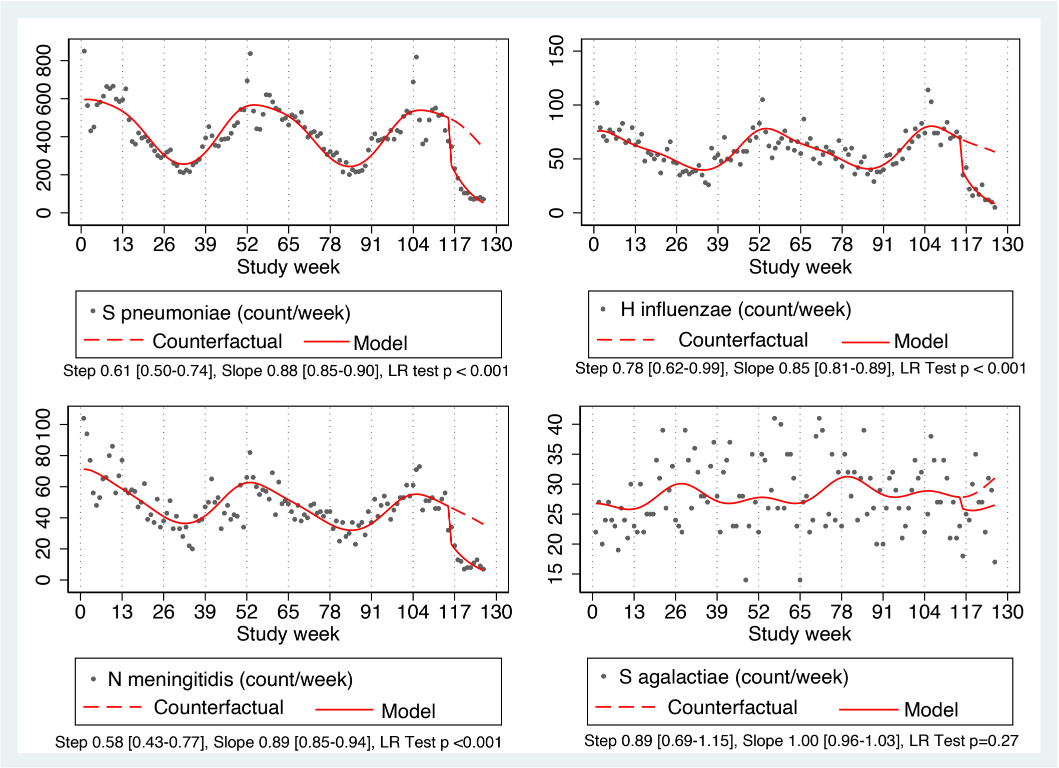


*Supplementary Figure 1. Observed count per week over the time period 1 January 2018 through 31 May 2020, a fitted model allowing for a step and slope change following week 11 of 2020 (study week 115), and a counterfactual model without this change for each species. Estimates for the step and slope (change per week) parameters and confidence intervals are given for each species, and a p-value from a likelihood ratio comparing models with both step and slope variables and models with neither.*

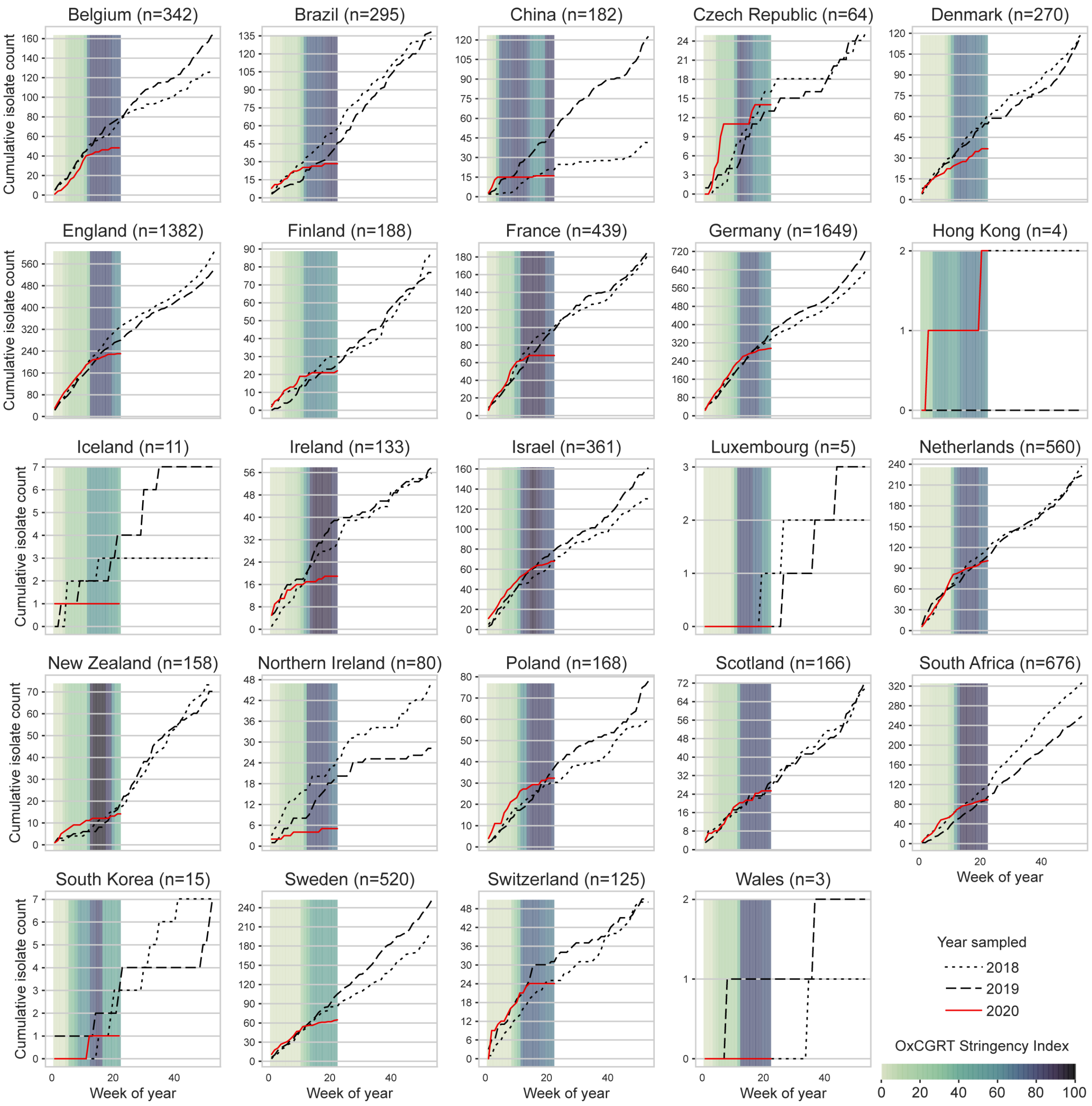


*Supplementary Figure 2. Annual cumulative curves of invasive H influenzae isolates submitted to IRIS laboratories in 23 countries from 1 January 2018 through 31 May 2020. Coloured bars represent the mean weekly OxCGRT Stringency Index values on a scale from 0-100. Larger (darker) values indicate that higher stringency measures were enacted within a country. Note that data from South Korea were submitted from two hospitals, one of which only started invasive disease surveillance in September 2018, so data presented here are only from then onwards.*

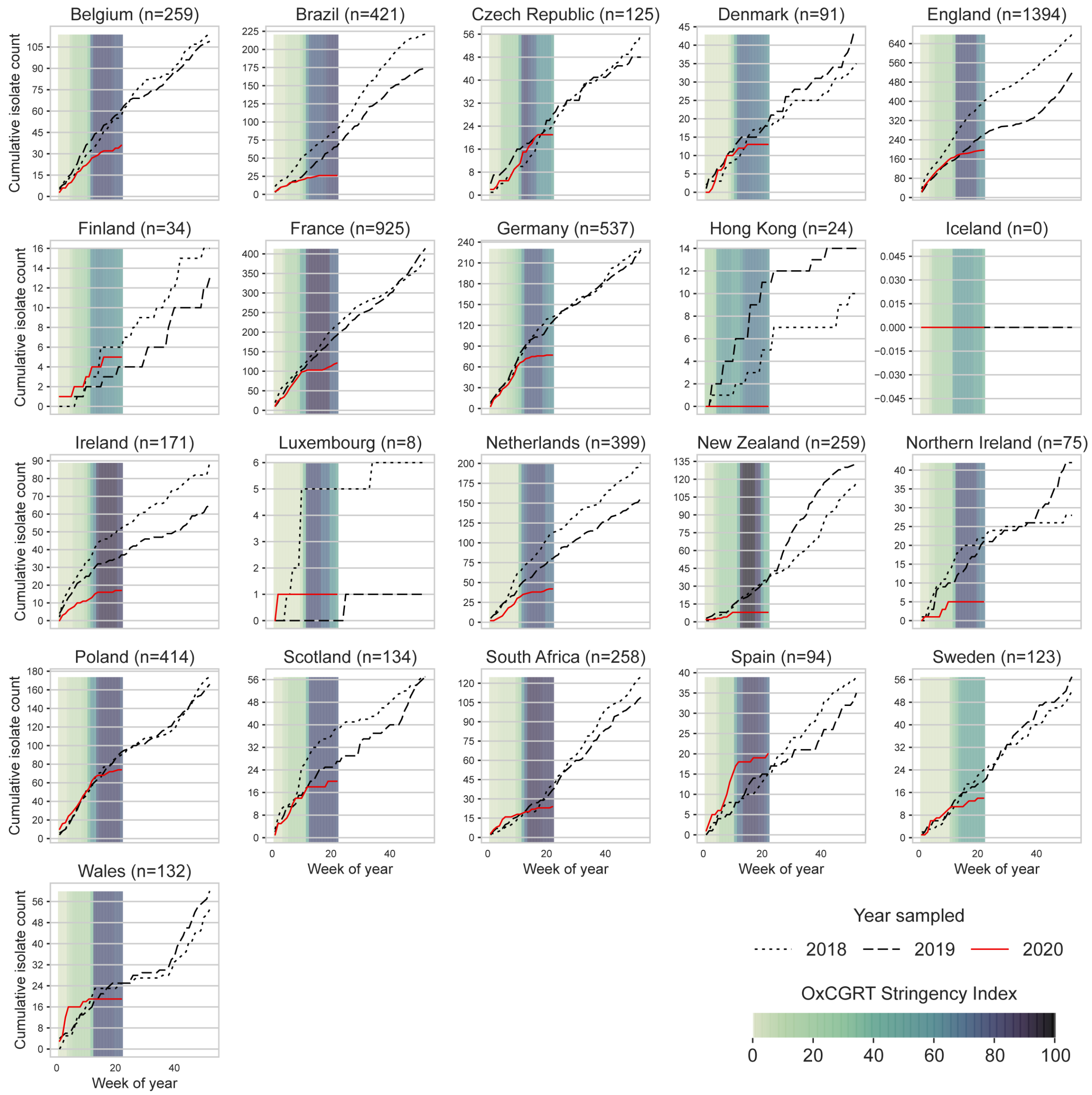


*Supplementary Figure 3. Annual cumulative curves of invasive N meningitidis isolates submitted to IRIS laboratories in 23 countries from 1 January 2018 through 31 May 2020. Coloured bars represent the mean weekly OxCGRT Stringency Index values on a scale from 0-100. Larger (darker) values indicate that higher stringency measures were enacted within a country.*

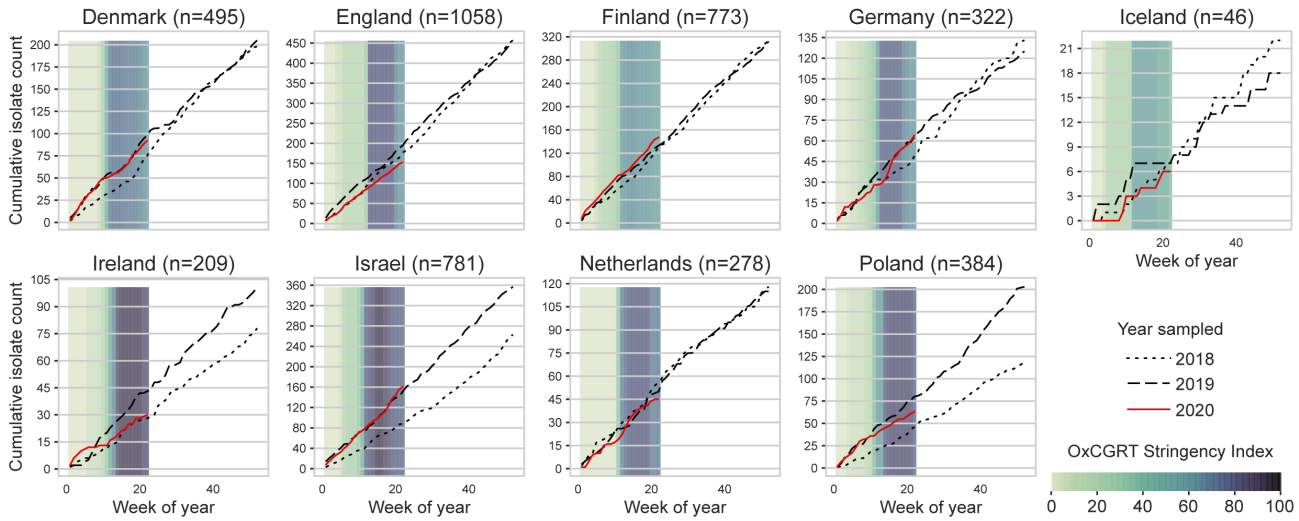


*Supplementary Figure 4. Annual cumulative curves of invasive S agalactiae isolates submitted to IRIS laboratories in nine countries from 1 January 2018 through 31 May 2020. Coloured bars represent the mean weekly OxCGRT Stringency Index values on a scale from 0-100. Larger (darker) values indicate that higher stringency measures were enacted within a country.*
